# A deep learning algorithm to detect cutaneous squamous cell carcinoma on frozen sections in Mohs micrographic surgery: a retrospective assessment

**DOI:** 10.1101/2023.05.14.23289960

**Authors:** Matthew J. Davis, Gokul Srinivasan, Rachael Chacko, Sophie Chen, Anish Suvarna, Louis J. Vaickus, Veronica C. Torres, Sassan Hodge, Eunice Y. Chen, Sarah Preum, Kimberley S. Samkoe, Brock C. Christensen, Matthew LeBoeuf, Joshua J. Levy

## Abstract

**Importance:** Intraoperative margin analysis is crucial for the successful removal of cutaneous squamous cell carcinomas (cSCC). Artificial intelligence technologies (AI) have previously demonstrated potential for facilitating rapid and complete tumor removal using intraoperative margin assessment for basal cell carcinoma. However, the varied morphologies of cSCC present challenges for AI margin assessment.

**Objective:** To develop and evaluate the accuracy of an AI algorithm for real-time histologic margin analysis of cSCC.

**Design:** A retrospective cohort study was conducted using frozen cSCC section slides and adjacent tissues.

**Setting:** This study was conducted in a tertiary care academic center.

**Participants:** Patients undergoing Mohs micrographic surgery for cSCC between January and March 2020.

**Exposures:** Frozen section slides were scanned and annotated, delineating benign tissue structures, inflammation, and tumor to develop an AI algorithm for real-time margin analysis. Patients were stratified by tumor differentiation status. Epithelial tissues including epidermis and hair follicles were annotated for moderate-well to well differentiated cSCC tumors. A convolutional neural network workflow was used to extract histomorphological features predictive of cSCC at 50-micron resolution.

**Main Outcomes and Measures:** The performance of the AI algorithm in identifying cSCC at 50-micron resolution was reported using the area under the receiver operating characteristic curve. Accuracy was also reported by tumor differentiation status and by delineation of cSCC from epidermis. Model performance using histomorphological features alone was compared to architectural features (i.e., tissue context) for well-differentiated tumors.

**Results:** The AI algorithm demonstrated proof of concept for identifying cSCC with high accuracy. Accuracy differed by differentiation status, driven by challenges in separating cSCC from epidermis using histomorphological features alone for well-differentiated tumors. Consideration of broader tissue context through architectural features improved the ability to delineate tumor from epidermis.

**Conclusions and Relevance:** Incorporating AI into the surgical workflow may improve efficiency and completeness of real-time margin assessment for cSCC removal, particularly in cases of moderately and poorly differentiated tumors/neoplasms. Further algorithmic improvement is necessary to remain sensitive to the unique epidermal landscape of well-differentiated tumors, and to map tumors to their original anatomical position/orientation. Future studies should assess the efficiency improvements and cost benefits and address other confounding pathologies such as inflammation and nuclei.

**Funding sources:** JL is supported by NIH grants R24GM141194, P20GM104416 and P20GM130454. Support for this work was also provided by the Prouty Dartmouth Cancer Center development funds.

**Key Points:** *Question:* How can the efficiency and accuracy of real-time intraoperative margin analysis for the removal of cutaneous squamous cell carcinoma (cSCC) be improved, and how can tumor differentiation be incorporated into this approach?

*Findings:* A proof-of-concept deep learning algorithm was trained, validated, and tested on frozen section whole slide images (WSI) for a retrospective cohort of cSCC cases, demonstrating high accuracy in identifying cSCC and related pathologies. Histomorphology alone was found to be insufficient to delineate tumor from epidermis in histologic identification of well-differentiated cSCC. Incorporation of surrounding tissue architecture and shape improved the ability to delineate tumor from normal tissue.

*Meaning:* Integrating artificial intelligence into surgical procedures has the potential to enhance the thoroughness and efficiency of intraoperative margin analysis for cSCC removal. However, accurately accounting for the epidermal tissue based on the tumor’s differentiation status requires specialized algorithms that consider the surrounding tissue context. To meaningfully integrate AI algorithms into clinical practice, further algorithmic refinement is needed, as well as the mapping of tumors to their original surgical site, and evaluation of the cost and efficacy of these approaches to address existing bottlenecks.

## Introduction

Cutaneous squamous cell carcinoma (cSCC) is the second most common form of skin cancer, with more than one million cases diagnosed in the United States each year ^1,2^. While many tumors are isolated to the skin, advanced disease is not uncommon with a metastasis rate of 4% and a disease-specific death estimate of 2.8% ^3^. When tumors occur on the head and neck or other high-risk sites, Mohs micrographic surgery (MMS) is the treatment of choice. MMS allows for real-time margin analysis resulting in low rates of recurrence. A recent study by Motley et al., found recurrence rates of cSCC of 3% when treated with MMS and 8% when treated with standard excision, despite a higher proportion of high risk tumors in the MMS group ^4^.

Early diagnosis and treatment of cutaneous tumors is essential. Currently, patient demand far outweighs the capacity of the dermatology workforce (Association of American Medical Colleges; AAMC), making early treatment more difficult ^5^. Machine learning models exist to detect basal cell carcinoma (BCC) ^5–7^, but given the complexities and variable morphologies of cSCC, similar algorithms are yet to be developed for this tumor type ^8^. Our study presents an algorithm to detect cSCC on whole slide images (WSI) of frozen sections obtained in MMS ^9^. Developing such algorithms may ameliorate the physician deficit by improving access to expert histologic assessment with the potential for broad application across numerous surgical specialties that treat various forms of cSCC.

## Methods

A retrospective study was designed, with WSI scanning (20X resolution using the Aperio AT2 image scanner) of 95 frozen section slides, each containing 3-5 tissue sections, followed by manual annotation of benign tissue structures, inflammation, and tumor by three dermatologists.

WSIs were then split into 256 × 256-pixel image patches (i.e., 50-micron resolution). Patches were randomly distributed into training, testing, and validation sets in an 80:10:10 arrangement, ensuring patches from the same patient were partitioned to the same set (e.g., restricting all patches across all tissue sections for one patient to the validation set). To classify tumors at the patch level, a convolutional neural network (CNN) workflow was implemented, using a ResNet101 model that was pre-trained and selected after comparing multiple neural network architectures (e.,g., SWIN-Transformer, EfficientNet) ^10^. The CNN workflow dynamically extracts histomorphological features at each 50-micron location, generating a probability score for cSCC between 0 and 1 ^11^. After the model was trained and validated, its performance characteristics were evaluated across the validation and testing sets using the Area Under the Receiver Operating Characteristic Curve (AUC), a performance metric that summarizes algorithmic sensitivity and specificity across a range of decision thresholds, with 95% confidence intervals reported using 1000-sample non-parametric bootstrapping.

Distinguishing cSCC from epithelial tissue based on histomorphology alone (i.e., what can be learned by a CNN) can be challenging, particularly in moderate-well to well differentiated squamous cell tumors, we hypothesized that the algorithm would not perform as well in these cases. To test this hypothesis, we annotated the epithelial tissue within the well-differentiated tumors in our cohort and compared the sensitivity and specificity of cSCC detection at 50-micron locations containing either cSCC or epithelium alone. To improve algorithmic performance in distinguishing cSCC from epithelium in well-differentiated tumors, we incorporated larger-scale architectural features beyond histomorphology. Specifically, we examined topological and shape descriptors, referred to as “architectural features”, of cSCC and epithelial tissue across the training, validation, and test sets ^12,13^. Topological and shape (i.e., architectural) features capture the relationships between tissue architectures and their shape properties. For instance, when viewed under a microscope, the epidermis typically appears flat or slightly curved, and may also have ridge-like features in certain areas; in contrast, cSCC is often characterized by a more discohesive and infiltrative growth pattern. The architectural features are numerical descriptors which encapsulate topological and shape differences and were used to train a random forest (RF) model for the purpose of distinguishing between SCC and epithelium. In addition, we incorporated a graph neural network (GNN) to consider contextual information from adjacent image patches ^14,15^. A graph neural network (GNN) is a type of neural network designed to operate on graphs and capture complex relationships and interactions between the nodes and edges of a graph. Unlike traditional neural networks that operate on vectorized inputs, GNNs can process structured data, which is useful for a variety of tasks such as node classification, link prediction, and graph clustering. For example, GNN increases the probability of classifying an image patch as epithelium if the surrounding patches were also classified as epithelial. We compared the performance of the architectural and GNN models to that of the CNN workflow to show how using the surrounding tissue architecture improves the accuracy of distinguishing SCC from epithelium in well differentiated tumors.

## Results

The algorithm achieved an AUC of 0.981 (95% CI [0.980-0.982]) and 0.935 (95% CI [0.934-0.936]) for predicting cSCC when applied to the validation and test sets respectively. As expected, the model performed better on poorly to moderately differentiated tumors (AUC=0.968, 95% CI [0.953-0.980]) than on well differentiated tumors (AUC=0.895, 95% CI [0.837-0.943]) (**Table 1, Figure 1, Supplementary Figures 1-3**). The difficulty in distinguishing normal epidermis from cSCC contributed to these deficiencies, yielding an AUC of 0.626 (95% CI [0.594-0.658]) when distinguishing cSCC from epithelium alone in well differentiated tumors (Figure 1). However, incorporating architectural (AUC=0.760; 95% CI [0.728-0.792]) and contextual (GNN; AUC=0.764; 95% CI [0.729-0.796]) features significantly improved the algorithm’s performance in delineating cSCC from epidermis (**Table 1, Figure 2**).

**Table 1:** Performance characteristics for SCC algorithm, considering histomorphological (CNN), architectural (topology/shape) and contextual (GNN) features across the validation and test sets, broken down by overall performance, tumor differentiation status, and restricting to SCC/epithelium within well-differentiated test set tumors; 95% confidence intervals reported using 1000-sample non-parametric bootstrapping

| Dataset | Algorithm | AUC | 2.5% CI | 97.5% CI |
| --- | --- | --- | --- | --- |
| **Validation Set: Overall** | **CNN/Morphology** | 0.981 | 0.980 | 0.982 |
| **Test Set: Overall** | **CNN/Morphology** | 0.935 | 0.934 | 0.936 |
| **Poor/Mod-Poor/Mod-Diff** | **CNN/Morphology** | 0.968 | 0.953 | 0.980 |
| **Mod-Well/Well-Diff** | **CNN/Morphology** | 0.895 | 0.837 | 0.943 |
| **cSCC versus Epidermis within Mod-Well/Well-Diff** | **CNN/Morphology** | 0.626 | 0.594 | 0.658 |
|  | **RF/Architecture** | 0.760 | 0.728 | 0.792 |
|  | **GNN/Context** | 0.764 | 0.729 | 0.796 |

**Figure 1:**
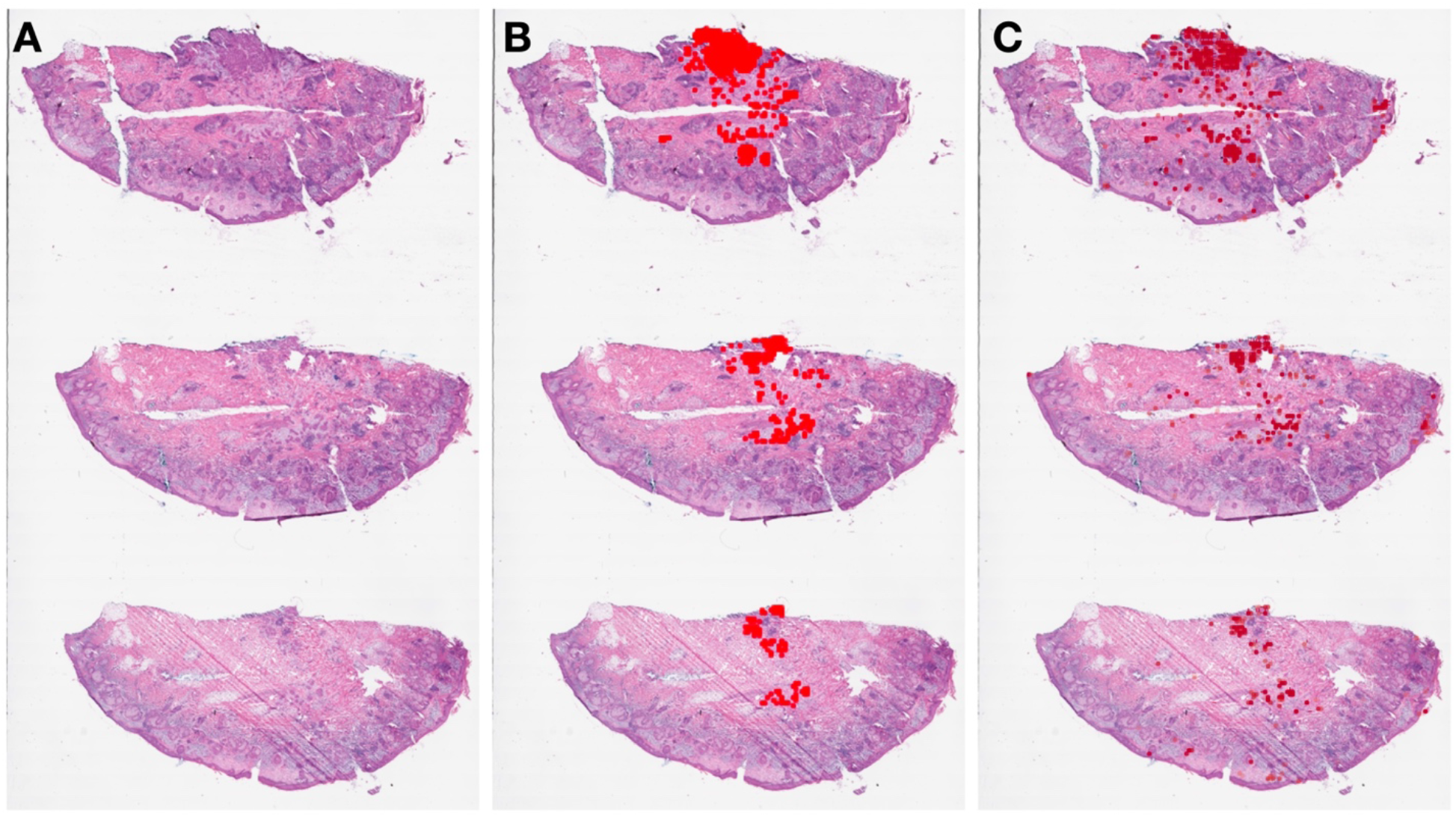
Example display output of cSCC prediction probabilities at 50-micron resolution for example test-set WSI: **A)** Original WSI; **B)** Ground truth cSCC; **C) c**SCC algorithm predictions.

**Figure 2:**
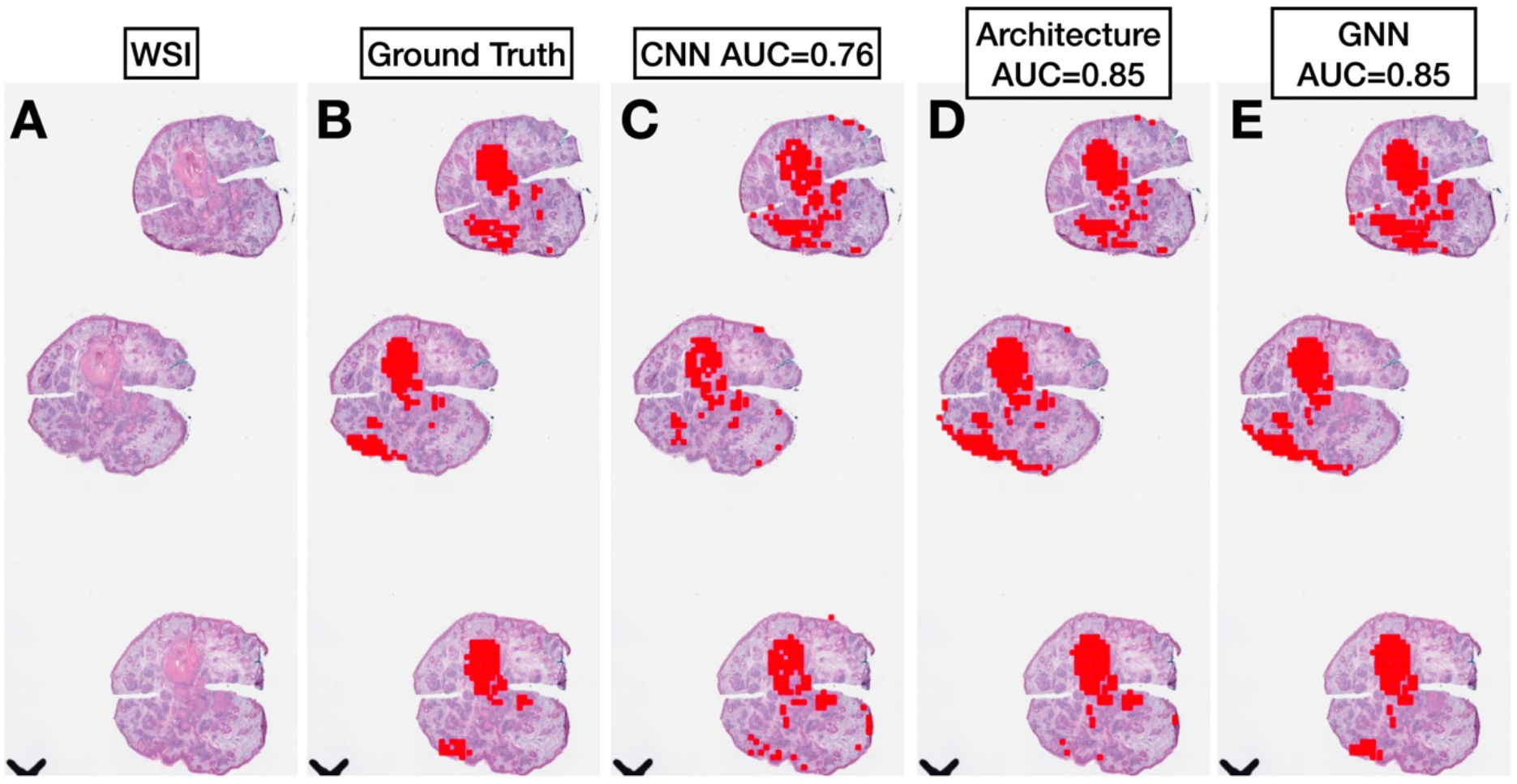
Example display output of cSCC prediction probabilities at 50-micron resolution for cSCC/Epithelium predictions across example test-set WSI for well-differentiated tumor: **A)** Original WSI; **B)** Ground truth cSCC; **C) c**SCC CNN algorithm predictions (histomorphology); **D)** Topological and shape features (architecture); **E)** GNN predictions (contextual)

## Discussion

Our study provides an example of a deep learning algorithm used to identify cutaneous squamous cell carcinoma on frozen section slides in MMS. Reducing rate limiting steps to intraoperative margin assessment of cSCC tumors can improve the efficiency and completeness of tumor removal, reducing the burden on laboratory staff while reducing tumor recurrence and repeat procedures ^16,17^. When evaluating this study, it should be acknowledged that all slides were obtained from a single MMS clinic and scanned images, not slides, were used for training, which may limit generalizability and real-world implementation. Application of this algorithm requires complete, high-quality tissue sections devoid of tears, holes, and other artifacts which may preclude histological margin assessment. Our data provide evidence supporting the identification of cSCC on frozen section slides, which has historically proven challenging. The algorithm’s successful performance in this study suggests its potential for broader use in providing real-time complete margin analysis of cSCC in various body parts. In the future, the focus will be on refining and improving the algorithm’s accuracy to enable more detailed identification of various associated pathologies, including single cell analysis, follicles, actinic keratosis, and incidental diagnoses. Additionally, efforts will be made to map tumors to their original anatomical position/orientation and evaluate the efficiency improvements and cost benefits of this algorithmic approach.

## Conclusion

This study not only established the general feasibility of histomorphological cSCC detection, but also demonstrated challenges in effectively distinguishing epithelium tissue from cSCC in well-differentiated cases. Therefore, algorithms that consider the surrounding tissue architecture could be useful for these tumors, although further research is necessary to improve the ability to utilize spatial cues. Furthermore, different tumor types may necessitate different algorithms. Future research will also address other confounding pathologies, such as inflammation, nuclei, follicles, architecture, and keratinocyte differentiation, by considering nuclei and large-scale architectural features.

## Data Availability

Due to patient privacy concerns, a subset data produced in the present study may be made available upon reasonable request to the authors.

## Acknowledgements

We would like to congratulate Sophie Chen for presenting an earlier iteration of this work at the national Junior Science and Humanities Symposium, placing 3^rd^ in the Computer Science competition.

## Supplementary Material

**Supplementary Figure 1:**
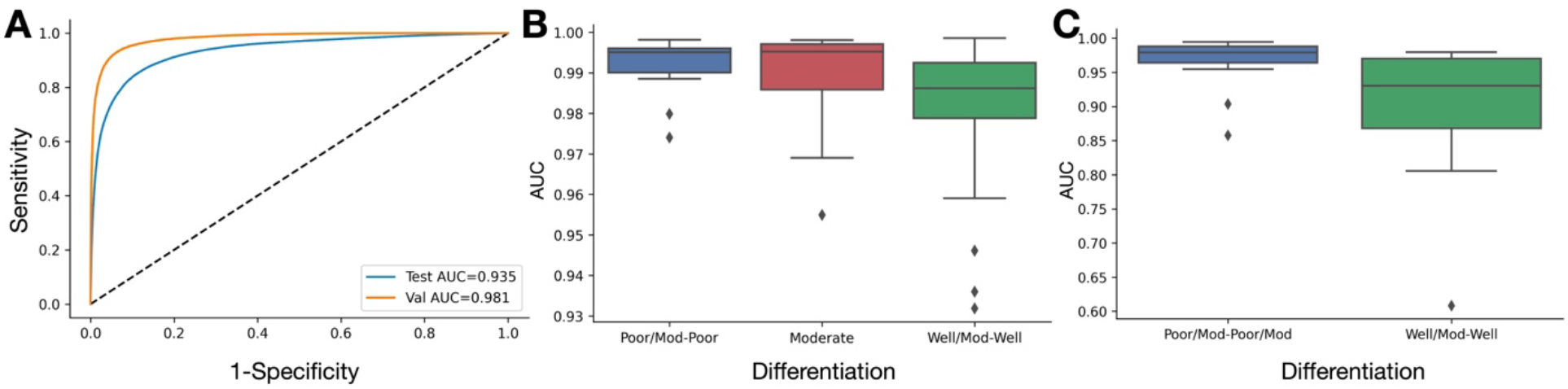
Performance characteristics of cSCC CNN algorithm: **A)** Receiver operating characteristic (ROC) curves for validation and test set cases at 50-micron resolution; **B)** Boxplots demonstrating distribution of slide-level AUC values from training set slides, broken down by differentiation status; **C)** Boxplots demonstrating distribution of slide-level AUC values from validation/test set slides, broken down by differentiation status

**Supplementary Figure 2:**
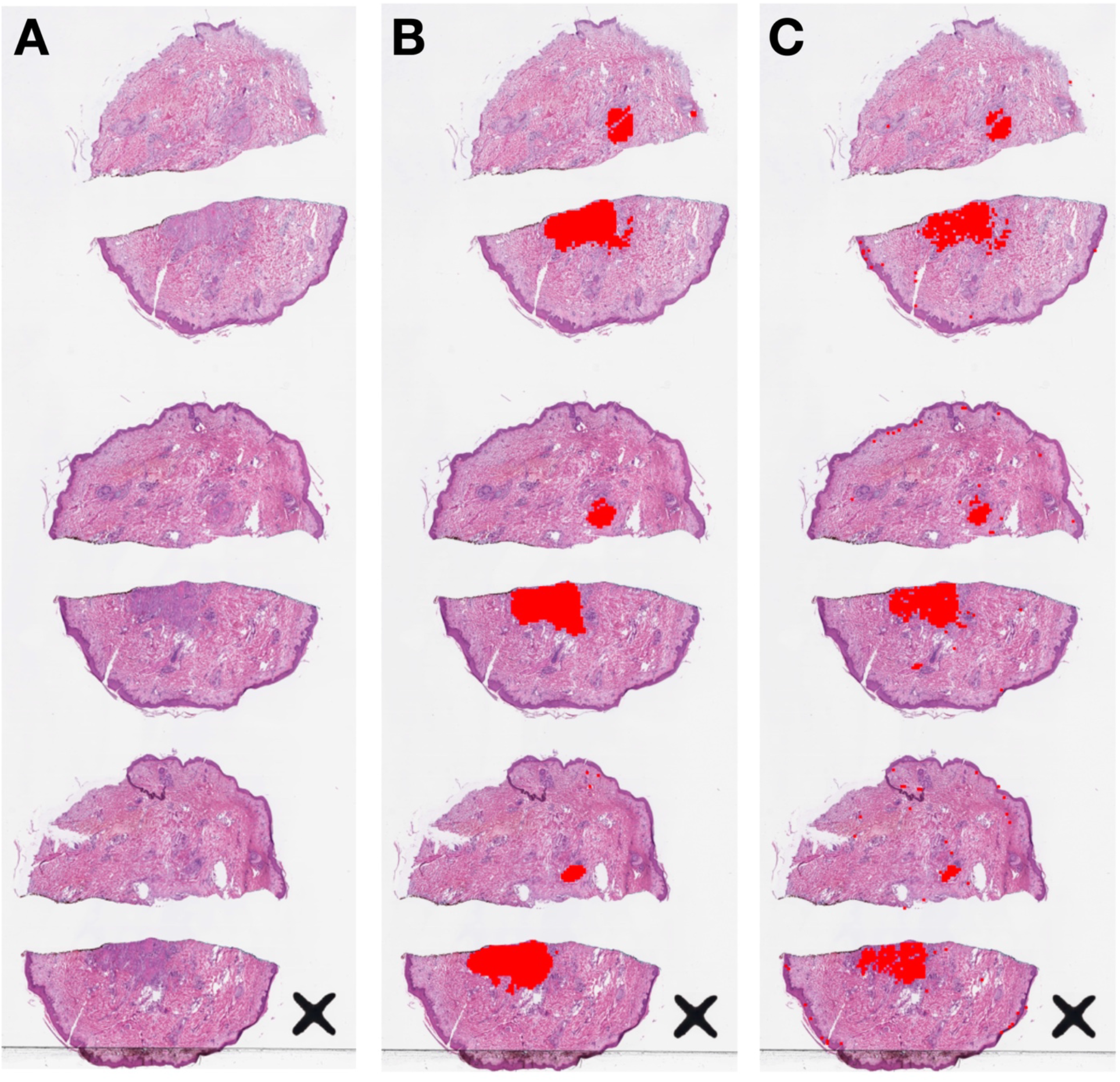
Example display output of sSCC prediction probabilities at 50-micron resolution for example validation-set WSI (poor-moderately differentiated): **A)** Original WSI; **B)** Ground truth cSCC; **C) c**SCC algorithm predictions

**Supplementary Figure 3:**
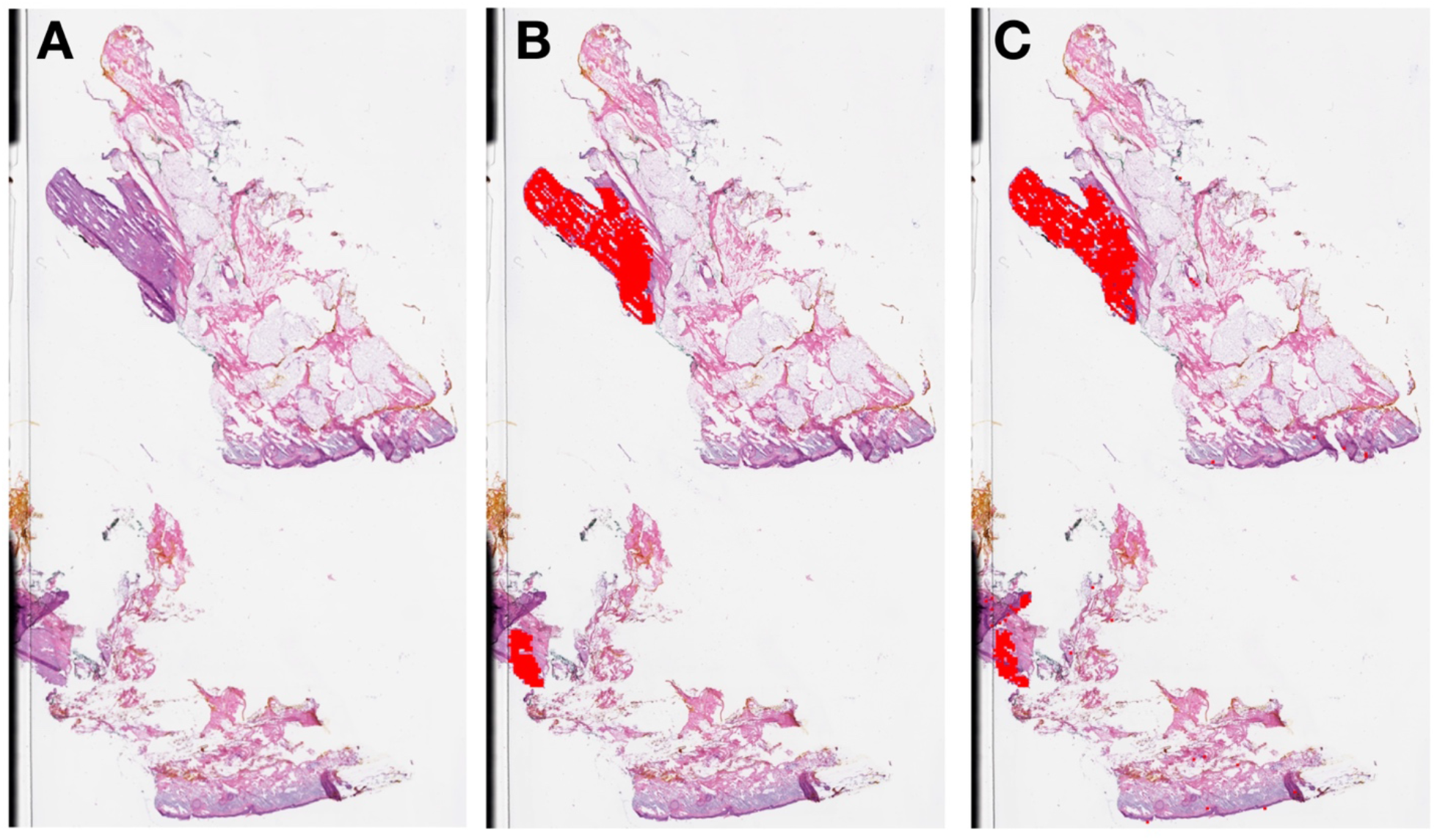
Example display output of cSCC prediction probabilities at 50-micron resolution for example validation-set WSI (poor-moderately differentiated): **A)** Original WSI; **B)** Ground truth cSCC; **C) c**SCC algorithm predictions

